# One-dose ChAdOx1 nCoV-19 Vaccine Effectiveness Against Symptomatic COVID-19 in a vulnerable community in Rio de Janeiro, Brazil: test-negative design study

**DOI:** 10.1101/2021.10.16.21265095

**Authors:** Otavio T Ranzani, Amanda A B Silva, Igor T Peres, Bianca B P Antunes, Thiago W Gonzaga-da-Silva, Daniel R Soranz, José Cerbino-Neto, Silvio Hamacher, Fernando A Bozza

## Abstract

We conducted a test-negative study design at the community “Complexo da Maré”, the largest group of favelas in Rio de Janeiro, Brazil, when Gamma and Delta were the predominant variants circulating. We estimated 42.4% (95% CI, 24.6, 56.0) protection against symptomatic COVID-19 after 21 days of one dose of ChAdOx1.

## INTRODUCTION

Several studies have reported the disproportionate impact COVID-19 is having on vulnerable communities [1–3]. This impact is due to the perpetuated social, economic and health inequities. Recently, inequity in access to vaccines has become a global issue [4]. Large populations in low- and middle-income countries (LMIC) live in slums or favelas, densely populated urban areas with deteriorated or incomplete infrastructure, but with a high risk of infectious disease transmission, limited access to health services or even vaccination.

Although the effectiveness of COVID-19 vaccines has been addressed in several articles from high income countries, studies that estimate vaccine effectiveness in neighborhoods such as favelas are lacking. We estimated the vaccine effectiveness of one dose of ChAdOx1 nCoV-19 (AstraZeneca/Oxford, hereafter ChAdOx1) vaccine against symptomatic COVID-19 using a test-negative design in the population of a large vulnerable community (“Complexo da Maré”) in Rio de Janeiro, Brazil.

## METHODS

“Complexo da Maré” is the largest group of favelas in Rio de Janeiro’s city, composed by 16 favelas with 140,000 residents [5], 54% of the population aged ≤30 years, and low HDI (0.686, 123rd out of Rio’s 126 neighbourhoods) in 2010 [6]. From the beginning of the pandemic until 14 September 2021, the region presented high rates of positive cases (6,416/100,000) and deaths (267/100,000) [7]. Since July 2020, a community broad testing strategy became available at the “Complexo da Maré” after an effort of civil society, NGOs and local community [8]. The testing was free of any charge and available on tents located in three different regions in Maré. There have been 193 RT-PCR tests per 1,000 inhabitants since the beginning of the campaign. During the study period, Gamma was the prevalent Variant of Concern in Rio de Janeiro and Delta became dominant after July 2021 [9]. This study was approved by the National Research Ethics Committee (IRB/CONEP) (CAAE - 49726921.6.0000.5248).

The COVID-19 vaccination campaign for Rio’s general population started on January 17, 2021, according to an age-based strategy. The vaccination campaign in “Complexo da Maré” initially followed the Rio’s city strategy, and by the end of July 2021, only 38% of Maré residents had received a first dose (53,084) and 13% a second or single dose (19,203) of COVID-19 vaccines. Then, Maré received a mass vaccination campaign, which applied about 36,000 first doses of ChAdOx1, over four days (July 29-August 1), achieving a coverage of about 85% of the adult population with at least 1 dose. Our analysis encompasses the period between Jan 17, 2021, and Sept 14, 2021. During this period, 136,026 doses (97,200 first doses and 38,826 second or single doses) were administered, being 83,762 doses (64,352 first doses and 19,410 second doses) of ChAdOx1. We did not analyze other vaccine platforms because of the small coverage proportion in the area.

We used a test-negative design to estimate the vaccine effectiveness of ChAdOx1 first dose, against symptomatic COVID-19. We linked the community-program testing database with the vaccination campaign database using a unique identifier (CPF, Cadastro de Pessoas Físicas). Overall, we followed the methodology reported elsewhere [10]. The description of inclusion/exclusion criteria is shown in eFigure 1. Briefly, we selected all RT-PCR (positive and negative tests) from symptomatic individuals, defined as presenting at least one symptom, from RT-PCRs sampled within 10 days of symptoms onset [11]. We excluded individuals with a previous positive RT-PCR, and those with a negative and subsequent positive test in the following 14 days. In a secondary analysis, we also included RT-PCR tests from asymptomatic individuals.

We estimated the vaccine effectiveness as 1-OR from logistic regression models. Our primary outcome was effectiveness against symptomatic COVID-19 after 21 days of the first dose of ChAdOx1. RT-PCR tests sampled after the second dose were excluded. We adjusted by time of epidemic using a restricted cubic spline on day of the year, and subsequently adjusted by age (restricted cubic spline), sex, self-reported race, Maré residence (three different regions - north, center and south), occupation, whether the RT-PCR was from routine testing or spontaneous walking, and for six chronic comorbidities. We evaluated the interaction between effectiveness and the age groups divided by the median of the symptomatic population (≤35 years; >35 years). We conducted four sensitivity analyses: 1) Excluding test-negative cases that reported taste/smell alterations [11]; 2) Expanding the time groups after the first dose until 56 days; 3) Analyzing symptomatic and asymptomatic cases, and 4) Analyzing only asymptomatic cases. We have missing data only for self-reported race (15%) and chronic comorbidities (<1%). We generated 20 multiple imputed datasets using the standard procedure for multiple imputation using chained equation. We summarized the estimates and generated 95% confidence intervals using the Rubin’s rules. All analyses were conducted in R statistical software version 4.0.3.

## RESULTS

Between Jan 17, 2021, and Sept 14, 2021, there were 15,771 RT-PCR tests in the community. After the inclusion/exclusion criteria (e-Figure 1), we analyzed 9,197 RT-PCR tests for the test-negative design. Overall, 36% of tests were from asymptomatic individuals. The test positivity was 20.6% (1211/5890) for symptomatic and 5.9% (195/3307) for asymptomatic cases (eFigure 2 and 3).

The general characteristics of the analyzed population for symptomatic cases is shown in eTable 1. The mean age was 38 ± 13 years, 65% of females and 41% of Brown/Pardo self-reported color. There were small proportion of individuals with chronic comorbidities, being most common obesity (9%) followed by cardiovascular disease (7%). Two-hundred and ten (3.6%) of individuals were healthcare workers. The median time between vaccination and RT-PCR testing among vaccinated was 36 [p25-p75: 20-60] days. The characteristics of the sample for asymptomatic and symptomatic and asymptomatic cases are shown in eTable 2 and eTable 3.

Vaccine effectiveness of one dose of ChAdOx1 is shown in Table 1. Fully adjusted vaccine effectiveness against symptomatic COVID-19 after 21 days of one dose was 42.4% (95% CI, 24.6%, 56.0%). The period 0-13 (bias-indicator) showed no indication of bias. For the sensitivity analysis for the primary outcome, excluding negative tests from individuals with taste/smell symptoms (n=4,951), the adjusted VE against symptomatic COVID-19 was 46.5% (95% CI, 29.4%, 59.5%) after 21 days (eTable 4).

**Table 1.**
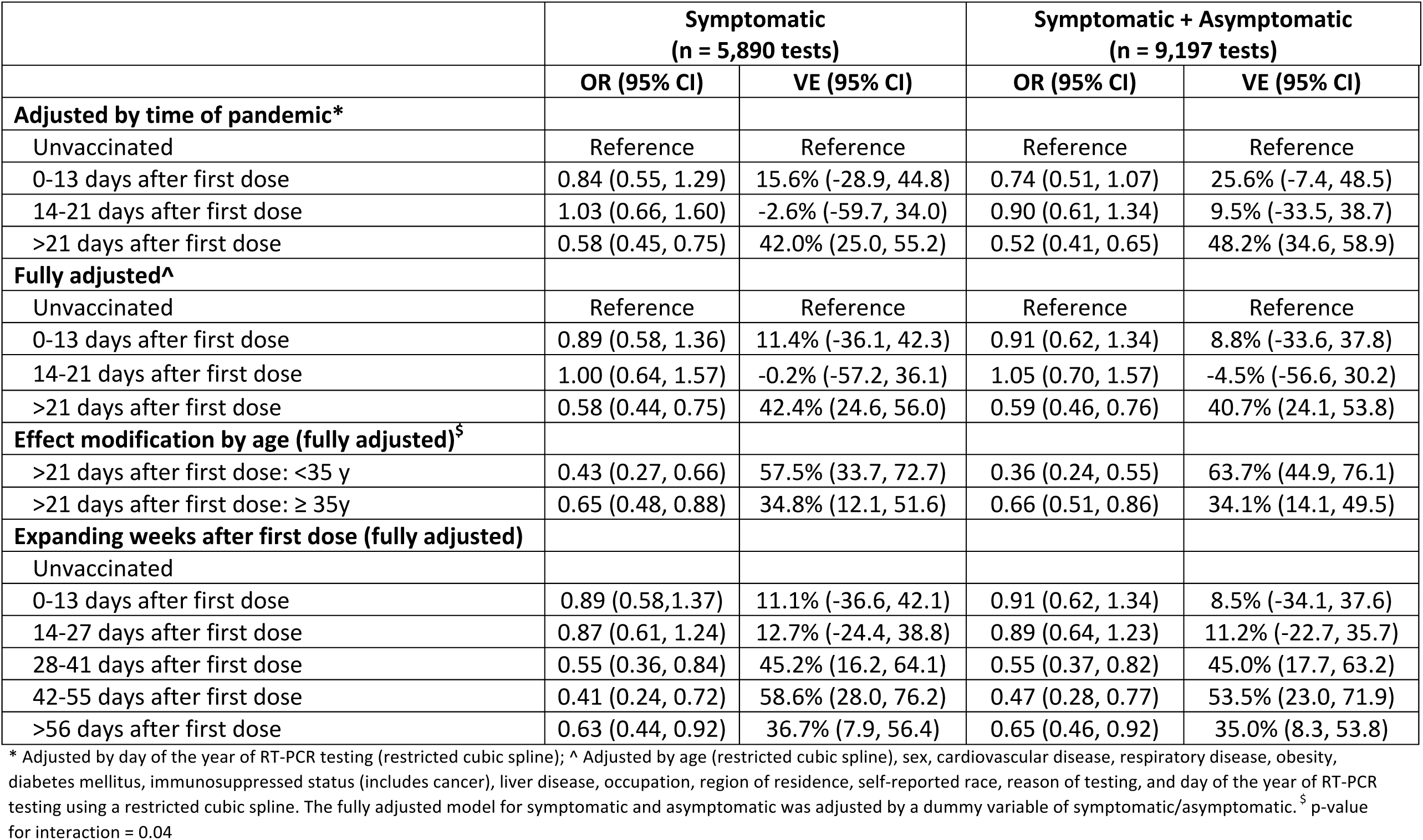
Vaccine effectiveness of the first dose of ChAdOx1

|  | Symptomatic<br>(n = 5,890 tests) |  | Symptomatic + Asymptomatic<br>(n = 9,197 tests) |  |
| --- | --- | --- | --- | --- |
|  | OR (95% CI) | VE (95% CI) | OR (95% CI) | VE (95% CI) |
| **Adjusted by time of pandemic\*** |  |  |  |  |
| Unvaccinated | Reference | Reference | Reference | Reference |
| 0-13 days after first dose | 0.84 (0.55, 1.29) | 15.6% (-28.9, 44.8) | 0.74 (0.51, 1.07) | 25.6% (-7.4, 48.5) |
| 14-21 days after first dose | 1.03 (0.66, 1.60) | -2.6% (-59.7, 34.0) | 0.90 (0.61, 1.34) | 9.5% (-33.5, 38.7) |
| >21 days after first dose | 0.58 (0.45, 0.75) | 42.0% (25.0, 55.2) | 0.52 (0.41, 0.65) | 48.2% (34.6, 58.9) |
| **Fully adjusted^** |  |  |  |  |
| Unvaccinated | Reference | Reference | Reference | Reference |
| 0-13 days after first dose | 0.89 (0.58, 1.36) | 11.4% (-36.1, 42.3) | 0.91 (0.62, 1.34) | 8.8% (-33.6, 37.8) |
| 14-21 days after first dose | 1.00 (0.64, 1.57) | -0.2% (-57.2, 36.1) | 1.05 (0.70, 1.57) | -4.5% (-56.6, 30.2) |
| >21 days after first dose | 0.58 (0.44, 0.75) | 42.4% (24.6, 56.0) | 0.59 (0.46, 0.76) | 40.7% (24.1, 53.8) |
| **Effect modification by age (fully adjusted)<sup>§</sup>** |  |  |  |  |
| >21 days after first dose: <35 y | 0.43 (0.27, 0.66) | 57.5% (33.7, 72.7) | 0.36 (0.24, 0.55) | 63.7% (44.9, 76.1) |
| >21 days after first dose: ≥ 35y | 0.65 (0.48, 0.88) | 34.8% (12.1, 51.6) | 0.66 (0.51, 0.86) | 34.1% (14.1, 49.5) |
| **Expanding weeks after first dose (fully adjusted)** |  |  |  |  |
| Unvaccinated |  |  |  |  |
| 0-13 days after first dose | 0.89 (0.58, 1.37) | 11.1% (-36.6, 42.1) | 0.91 (0.62, 1.34) | 8.5% (-34.1, 37.6) |
| 14-27 days after first dose | 0.87 (0.61, 1.24) | 12.7% (-24.4, 38.8) | 0.89 (0.64, 1.23) | 11.2% (-22.7, 35.7) |
| 28-41 days after first dose | 0.55 (0.36, 0.84) | 45.2% (16.2, 64.1) | 0.55 (0.37, 0.82) | 45.0% (17.7, 63.2) |
| 42-55 days after first dose | 0.41 (0.24, 0.72) | 58.6% (28.0, 76.2) | 0.47 (0.28, 0.77) | 53.5% (23.0, 71.9) |
| >56 days after first dose | 0.63 (0.44, 0.92) | 36.7% (7.9, 56.4) | 0.65 (0.46, 0.92) | 35.0% (8.3, 53.8) |

\* Adjusted by day of the year of RT-PCR testing (restricted cubic spline); ^ Adjusted by age (restricted cubic spline), sex, cardiovascular disease, respiratory disease, obesity, diabetes mellitus, immunosuppressed status (includes cancer), liver disease, occupation, region of residence, self-reported race, reason of testing, and day of the year of RT-PCR testing using a restricted cubic spline. The fully adjusted model for symptomatic and asymptomatic was adjusted by a dummy variable of symptomatic/asymptomatic. <sup>§</sup> p-value for interaction = 0.04

There was an interaction for age groups, with the young group (age ≤35 years) showing high effectiveness (57.5%; 95% CI, 33.7%, 72.7%; P_interaction_ = 0.04). The adjusted VE increased for subsequent days after the first dose, except for >56 days. The vaccine effectiveness when considering those symptomatic and asymptomatic together (40.7%; 95% CI, 24.1%, 53.8%) was comparable to the main analysis. The vaccine effectiveness only among asymptomatic tests was 29.8% (95% CI, -44.2%, 65.8%, eTable 5).

## DISCUSSION

We observed a vaccine effectiveness of 42% against symptomatic COVID-19 after a single dose of ChAdOx1 in a vulnerable population in Rio de Janeiro, Brazil, in a period of mixed Gamma and Delta dominance.

Our estimate is in accordance with previous evaluations for vaccine effectiveness of the first dose of ChAdOx1 in the context of Gamma or Delta [12,13]. We observed the vaccine effectiveness increased up to 58.6% (28.0, 76.2) during 42-55 days of the first dose, which has been reported [12] and a decrease afterwards. We can hypothesize that this decrease might occur because of some factors, such limited power to evaluate vaccine effectiveness at this time window and increase on the dominance of Delta.

There is limited evidence for vaccine effectiveness against infection. We aimed to evaluate it combining asymptomatic cases in the estimates. The estimates are comparable to the protection against symptomatic cases, as previously reported [13]. However, the limited sample size resulting in low number of events among asymptomatic cases, shifts the estimate of the combined analysis towards symptomatic cases. Our analyses only with asymptomatic cases, although with large imprecision, shows a potential for effectiveness against asymptomatic infection. A detailed analysis on symptoms, such as a follow-up on those asymptomatic at RT-PCR collection or active surveys regarding symptoms, could help on understanding vaccine effectiveness against infection.

Our study has some limitations. First, we could not evaluate second dose effectiveness and waning because of the vaccination campaign and enough follow-up. Second, data on genome sequencing from all test-positives was not available. However, our study analyzed data in a period of high transmission rates of the Gamma and Delta variants in the community. Third, we could not evaluate the vaccine effectiveness for preventing COVID-19 hospitalizations or severe outcomes. Finally, although we excluded previous confirmed infections, it is expected that the community has a high attack rate and so the unvaccinated could have an unmeasured protection level, underestimating the vaccine effectiveness.

One-dose of ChAdOx1 was effective on reducing symptomatic COVID-19 in using data from a community surveillance program, where there was a broad testing strategy without any cost to an overall young vulnerable population in a group of favelas in Brazil.

## Data Availability

All data produced in the present study are available upon reasonable request to the authors.

## Acknowledgments

We thank the Redes da Mare for all support and the efficient strategies on community engagement and communication during the pandemic and the Unidade de Apoio ao Diagnóstico da Covid-19 (UNADIG-FIOCRUZ) for the support on the testing diagnosis. OTR acknowledges support from the Spanish Ministry of Science and Innovation through the Centro de Excelencia Severo Ochoa 2019-2023 Program and from the Generalitat de Catalunya through the CERCA Program. We thank the Center for Healthcare Operations and Intelligence (NOIS) research group for their discussions and collaborative production of scientific analyses of the COVID-19 pandemic in Brazil.

## Supplementary Data

**eFigure 1.**
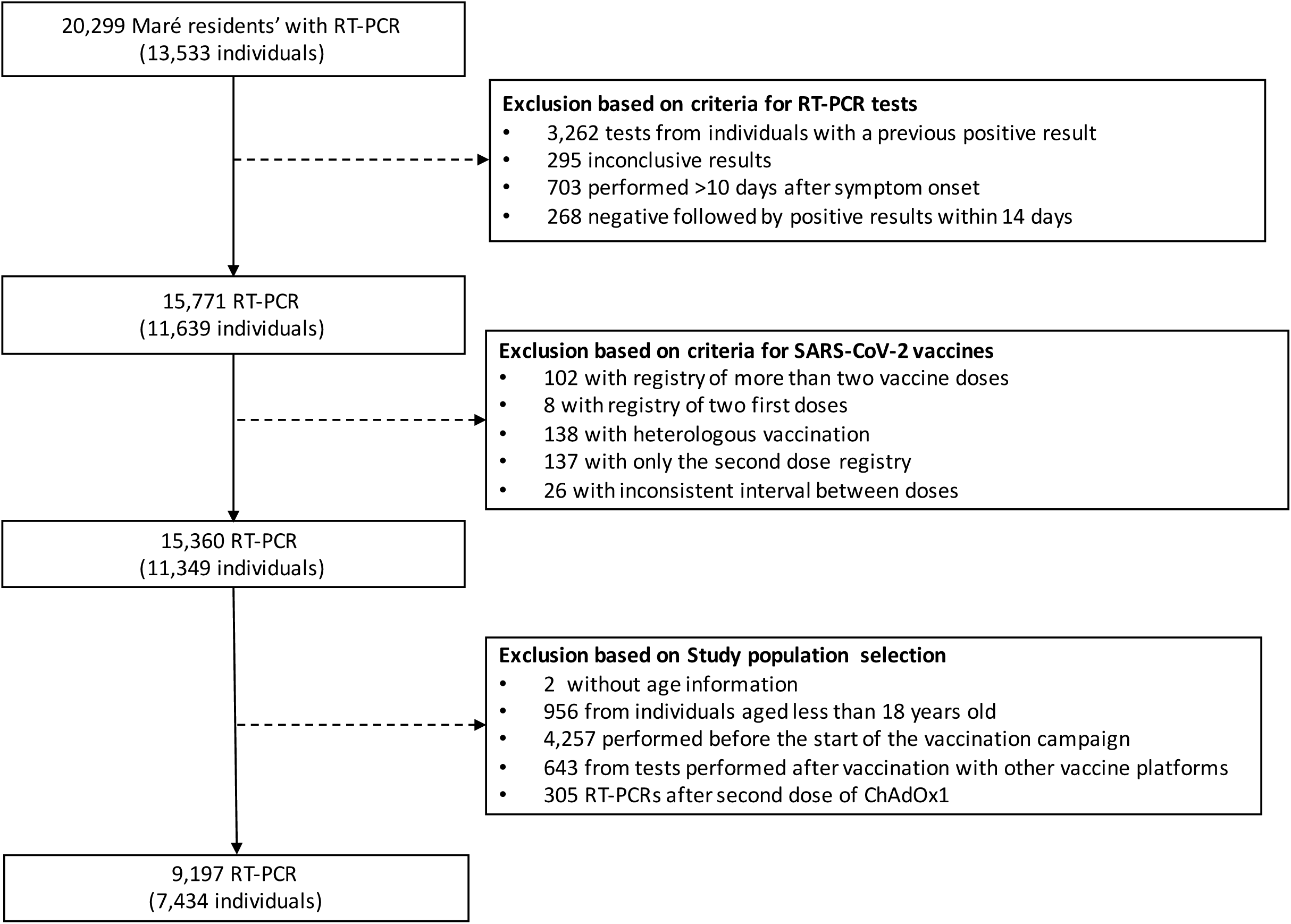
Flowchart.

**eFigure 2.**
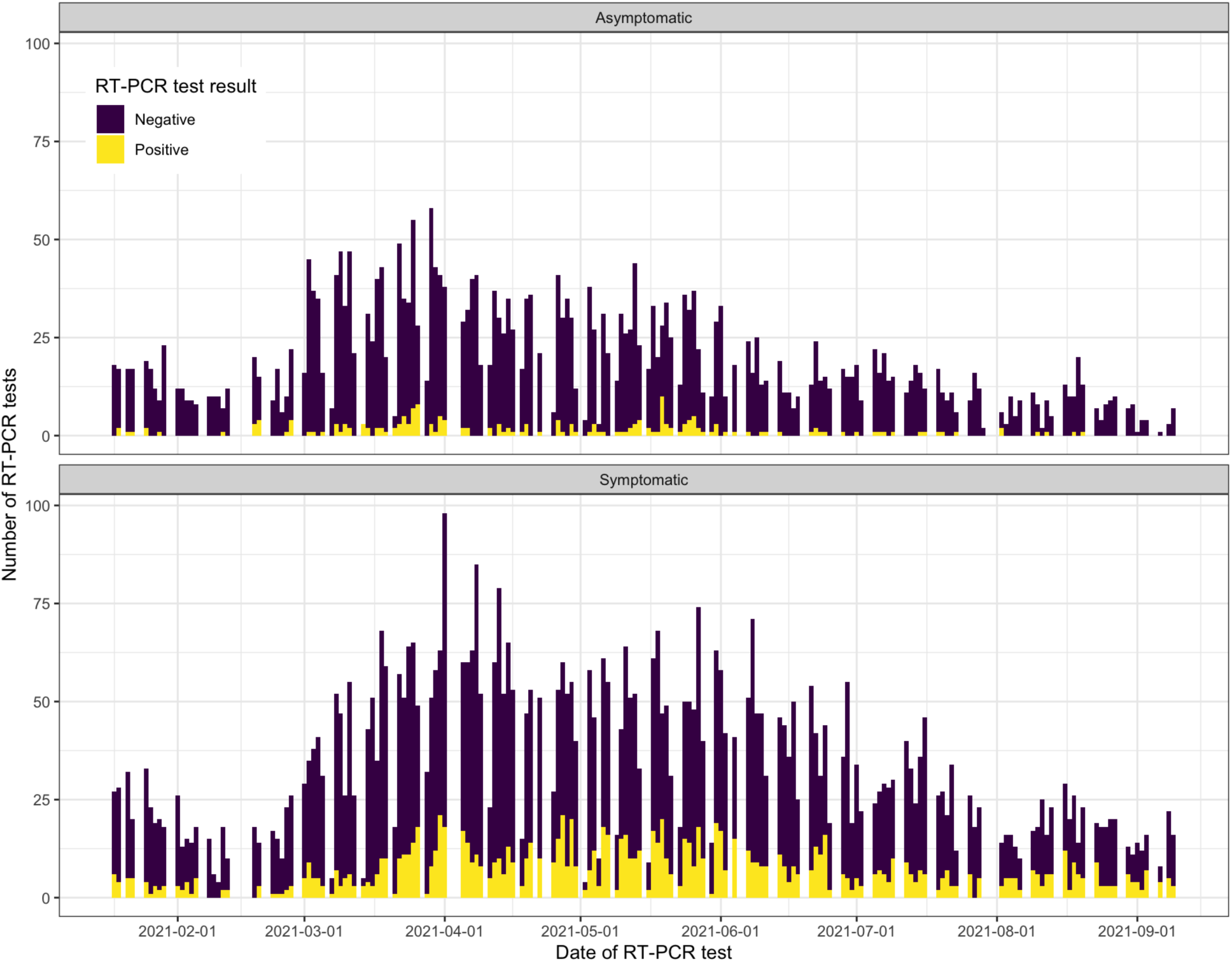
RT-PCR tests and results during the study period stratified by presence of symptoms.

**eFigure3.**
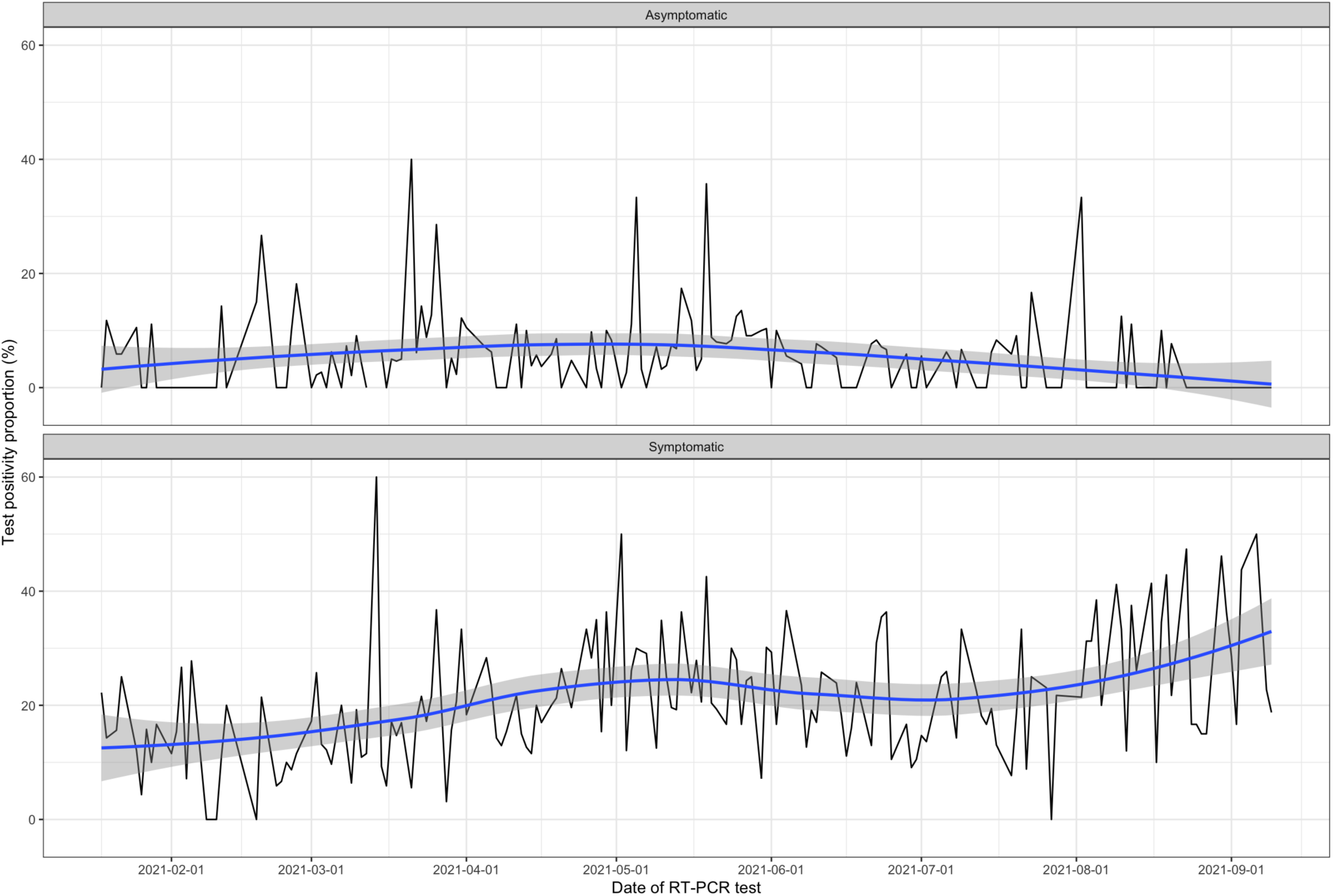
Test positivity proportion during the study period stratified by presence of symptoms. The blue line represents the smooth curve and shade area the 95% confidence intervals

**eFigure 4.**
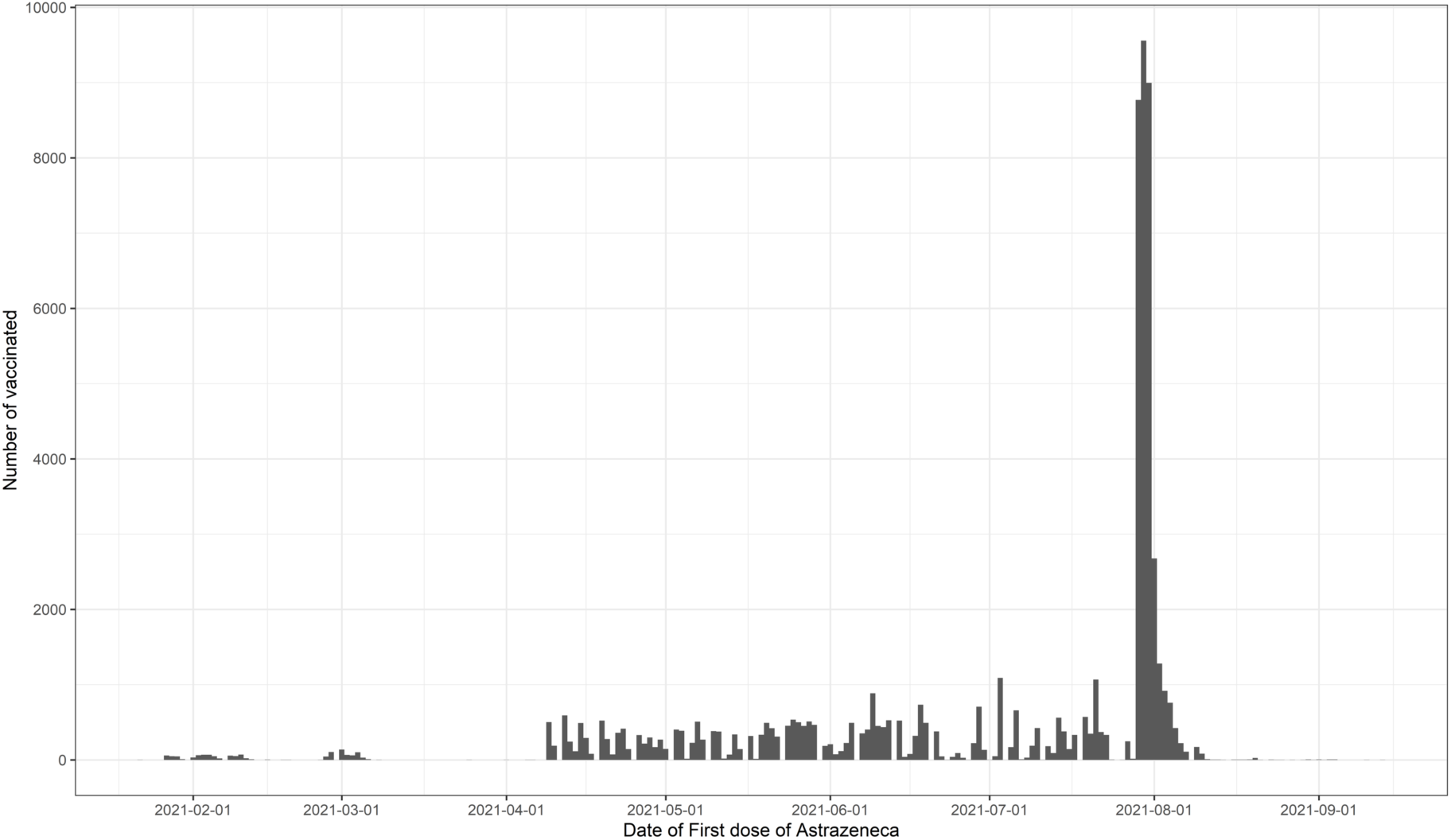
First doses of AstraZeneca vaccine during the vaccination campaign in the “Complexo da Maré”.

**eTable 1.** Characteristics for the symptomatic cases.

|  | Overall<br>(n=5,890) | Test Negative<br>(n=4679) | Test Positive<br>(n=1211) |
| --- | --- | --- | --- |
| Age, mean ± SD | 38.0 (13) | 38.0 (13) | 38.1 (13) |
| Age ≤35 years | 2838 (48.2) | 2249 (48.1) | 589 (48.6) |
| Age >35 years | 3052 (51.8) | 2430 (51.9) | 622 (51.4) |
| **Sex** |  |  |  |
| Female | 3809 (64.7) | 3079 (65.8) | 730 (60.3) |
| Male | 2081 (35.3) | 1600 (34.2) | 481 (39.7) |
| **Self-reported race** |  |  |  |
| White | 1443 (24.5) | 1149 (24.6) | 294 (24.3) |
| Brown | 2383 (40.5) | 1902 (40.6) | 481 (39.7) |
| Black | 1021 (17.3) | 822 (17.6) | 199 (16.4) |
| Other | 143 (2.4) | 116 (2.5) | 27 (2.2) |
| Missing | 900 (15.3) | 690 (14.7) | 210 (17.3) |
| **Comorbidities\*** |  |  |  |
| Cardiovascular disease | 420 (7.1) | 350 (7.5) | 70 (5.8) |
| Respiratory disease | 182 (3.1) | 153 (3.3) | 29 (2.4) |
| Immunosuppressed status | 231 (3.9) | 190 (4.1) | 41 (3.4) |
| Liver disease | 97 (1.6) | 76 (1.6) | 21 (1.7) |
| Obesity | 536 (9.1) | 450 (9.6) | 86 (7.1) |
| Diabetes mellitus | 393 (6.7) | 327 (7.0) | 66 (5.5) |
| **Occupation** |  |  |  |
| Other | 5613 (95.3) | 4437 (94.8) | 1176 (97.1) |
| Security | 67 (1.1) | 56 (1.2) | 11 (0.9) |
| Healthcare worker | 210 (3.6) | 186 (4.0) | 24 (2.0) |
| **Region of residence** |  |  |  |
| Centre | 3164 (53.7) | 2555 (54.6) | 609 (50.3) |
| North | 417 (7.1) | 324 (6.9) | 93 (7.7) |
| South | 2309 (39.2) | 1800 (38.5) | 509 (42.0) |
| **Vaccination status** |  |  |  |
| **Main analysis** |  |  |  |
| Unvaccinated | 4988 (84.7) | 3956 (84.5) | 1032 (85.2) |
| 0-13 days after first dose | 132 (2.2) | 102 (2.2) | 30 (2.5) |
| 14-21 days after first dose | 111 (1.9) | 82 (1.8) | 29 (2.4) |
| >21 days after first dose | 659 (11.2) | 539 (11.5) | 120 (9.9) |
| **Sensitivity analysis** |  |  |  |
| Unvaccinated | 4988 (84.7) | 3956 (84.5) | 1032 (85.2) |
| 0-13 days after first dose | 132 (2.2) | 102 (2.2) | 30 (2.5) |
| 14-27 days after first dose | 204 (3.5) | 156 (3.3) | 48 (4.0) |
| 28-41 days after first dose | 182 (3.1) | 149 (3.2) | 33 (2.7) |
| 42-55 days after first dose | 127 (2.2) | 111 (2.4) | 16 (1.3) |
| >56 days after first dose | 257 (4.4) | 205 (4.4) | 52 (4.3) |
| **Time between vaccination and PCR, median [p25, p75]** | 36 [20, 60] | 37 [20, 59] | 32 [18, 61] |

\* 2 missing values for each comorbidity. Data as N (%) unless differently reported

**eTable 2.** Characteristics for the asymptomatic cases.

|  | Overall<br>(n=3307) | Test Negative<br>(n=3112) | Test Positive<br>(n=195) |
| --- | --- | --- | --- |
| Age, mean ± SD | 42.6 (15) | 42.6 (15) | 42.4 (15) |
| Age ≤35 years | 1161 (35.1) | 1091 (35.1) | 70 (35.9) |
| Age >35 years | 2146 (64.9) | 2021 (64.9) | 125 (64.1) |
| **Sex** |  |  |  |
| Female | 1952 (59.0) | 1834 (58.9) | 118 (60.5) |
| Male | 1355 (41.0) | 1278 (41.1) | 77 (39.5) |
| **Self-reported race** |  |  |  |
| White | 607 (18.4) | 571 (18.3) | 36 (18.5) |
| Brown/Pardo | 1116 (33.7) | 1048 (33.7) | 68 (34.9) |
| Black | 667 (20.2) | 628 (20.2) | 39 (20.0) |
| Other | 95 (2.9) | 90 (2.9) | 5 (2.6) |
| Missing | 822 (24.9) | 775 (24.9) | 47 (24.1) |
| **Comorbidities\*** |  |  |  |
| Cardiovascular disease | 254 (7.7) | 242 (7.8) | 12 (6.2) |
| Respiratory disease | 41 (1.2) | 38 (1.2) | 3 (1.5) |
| Imunossupressed status | 66 (2.0) | 63 (2.0) | 3 (1.5) |
| Liver disease | 23 (0.7) | 23 (0.7) | 0 (0.0) |
| Obesity | 159 (4.8) | 154 (4.9) | 5 (2.6) |
| Diabetes mellitus | 190 (5.7) | 185 (5.9) | 5 (2.6) |
| **Occupation** |  |  |  |
| Other | 3180 (96.2) | 2988 (96.0) | 192 (98.5) |
| Security | 38 (1.1) | 37 (1.2) | 1 (0.5) |
| Healthcare worker | 89 (2.7) | 87 (2.8) | 2 (1.0) |
| **Region of residence** |  |  |  |
| Centre | 1765 (53.4) | 1664 (53.5) | 101 (51.8) |
| North | 305 (9.2) | 288 (9.3) | 17 (8.7) |
| South | 1237 (37.4) | 1160 (37.3) | 77 (39.5) |
| **Vaccination status** |  |  |  |
| **Main analysis** |  |  |  |
| Unvaccinated | 2731 (82.6) | 2559 (82.2) | 172 (88.2) |
| 0-13 days after first dose | 102 (3.1) | 96 (3.1) | 6 (3.1) |
| 14-21 days after first dose | 72 (2.2) | 67 (2.2) | 5 (2.6) |
| >21 days after first dose | 402 (12.2) | 390 (12.5) | 12 (6.2) |
| **Sensitivity analysis** |  |  |  |
| Unvaccinated | 2731 (82.6) | 2559 (82.2) | 172 (88.2) |
| 0-13 days after first dose | 102 (3.1) | 96 (3.1) | 6 (3.1) |
| 14-27 days after first dose | 135 (4.1) | 128 (4.1) | 7 (3.6) |
| 28-41 days after first dose | 88 (2.7) | 87 (2.8) | 1 (0.5) |
| 42-55 days after first dose | 72 (2.2) | 69 (2.2) | 3 (1.5) |
| >56 days after first dose | 179 (5.4) | 173 (5.6) | 6 (3.1) |
| **Time between vaccination and PCR, median [p25, p75]** | 35 [18, 61] | 35 [18, 61] | 24 [13, 56] |

\* 1 missing value for each comorbidity. Data as N (%) unless differently reported

**eTable 3.** Characteristics for symptomatic and asymptomatic cases.

|  | Overall<br>(n=9197) | Test Negative<br>(n=7791) | Test Positive<br>(n=1406) |
| --- | --- | --- | --- |
| Age, mean ± SD | 39.6 (14) | 39.8 (14) | 38.7 (14) |
| Age ≤35 years | 3999 (43.5) | 3340 (42.9) | 659 (46.9) |
| Age >35 years | 5198 (56.5) | 4451 (57.1) | 747 (53.1) |
| **Sex** |  |  |  |
| Female | 5761 (62.6) | 4913 (63.1) | 848 (60.3) |
| Male | 3436 (37.4) | 2878 (36.9) | 558 (39.7) |
| **Self-reported race** |  |  |  |
| White | 2050 (22.3) | 1720 (22.1) | 330 (23.5) |
| Brown | 3499 (38.0) | 2950 (37.9) | 549 (39.0) |
| Black | 1688 (18.4) | 1450 (18.6) | 238 (16.9) |
| Other | 238 (2.6) | 206 (2.6) | 32 (2.3) |
| Missing | 1722 (18.7) | 1465 (18.8) | 257 (18.3) |
| **Comorbidities\*** |  |  |  |
| Cardiovascular disease | 674 (7.3) | 592 (7.6) | 82 (5.8) |
| Respiratory disease | 223 (2.4) | 191 (2.5) | 32 (2.3) |
| Immunosuppressed status | 297 (3.2) | 253 (3.2) | 44 (3.1) |
| Liver disease | 120 (1.3) | 99 (1.3) | 21 (1.5) |
| Obesity | 695 (7.6) | 604 (7.8) | 91 (6.5) |
| Diabetes mellitus | 583 (6.3) | 512 (6.6) | 71 (5.0) |
| **Occupation** |  |  |  |
| Other | 8793 (95.6) | 7425 (95.3) | 1368 (97.3) |
| Security | 105 (1.1) | 93 (1.2) | 12 (0.9) |
| Healthcare worker | 299 (3.3) | 273 (3.5) | 26 (1.8) |
| **Region of residence** |  |  |  |
| Centre | 4929 (53.6) | 4219 (54.2) | 710 (50.5) |
| North | 722 (7.9) | 612 (7.9) | 110 (7.8) |
| South | 3546 (38.6) | 2960 (38.0) | 586 (41.7) |
| **Vaccination status** |  |  |  |
| **Main analysis** |  |  |  |
| Unvaccinated | 7719 (83.9) | 6515 (83.6) | 1204 (85.6) |
| 0-13 days after first dose | 234 (2.5) | 198 (2.5) | 36 (2.6) |
| 14-21 days after first dose | 183 (2.0) | 149 (1.9) | 34 (2.4) |
| >21 days after first dose | 1061 (11.5) | 929 (11.9) | 132 (9.4) |
| **Sensitivity analysis** |  |  |  |
| Unvaccinated | 7719 (83.9) | 6515 (83.6) | 1204 (85.6) |
| 0-13 days after first dose | 234 (2.5) | 198 (2.5) | 36 (2.6) |
| 14-27 days after first dose | 339 (3.7) | 284 (3.6) | 55 (3.9) |
| 28-41 days after first dose | 270 (2.9) | 236 (3.0) | 34 (2.4) |
| 42-55 days after first dose | 199 (2.2) | 180 (2.3) | 19 (1.4) |
| >56 days after first dose | 436 (4.7) | 378 (4.9) | 58 (4.1) |
| **Time between vaccination and PCR, median [p25, p75]** | 36 [19, 60] | 36 [20, 60] | 32 [17, 60] |

\* 5 missing values for each comorbidity. Data as N (%) unless differently reported

**eTable 4.**
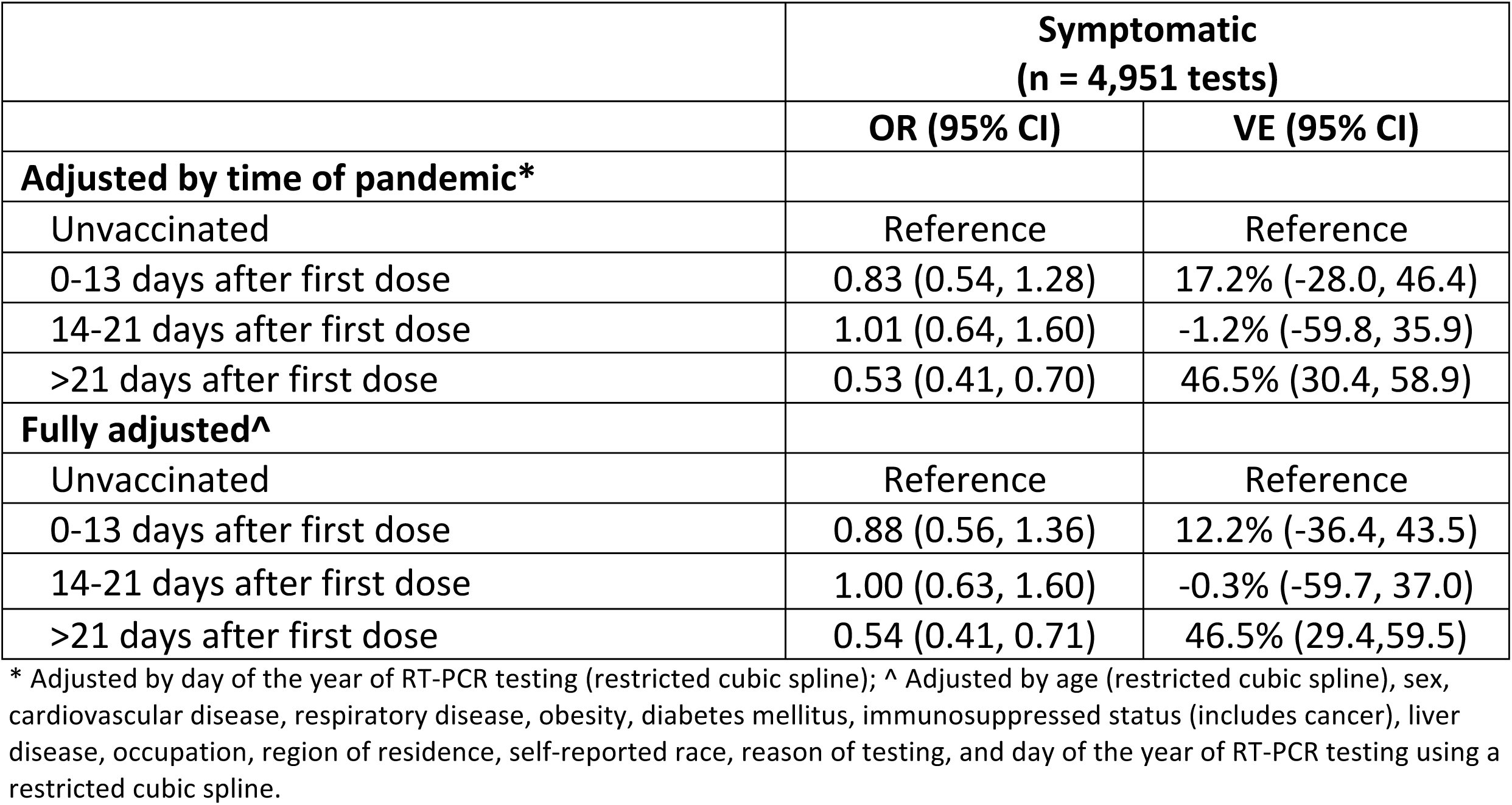
Vaccine effectiveness against symptomatic COVID-19. (sensitivity analysis excluding test-negatives with taste/smell symptoms)

|  | **Symptomatic**<br>**(n = 4,951 tests)** |  |
| --- | --- | --- |
|  | **OR (95% CI)** | **VE (95% CI)** |
| **Adjusted by time of pandemic\*** |  |  |
| Unvaccinated | Reference | Reference |
| 0-13 days after first dose | 0.83 (0.54, 1.28) | 17.2% (-28.0, 46.4) |
| 14-21 days after first dose | 1.01 (0.64, 1.60) | -1.2% (-59.8, 35.9) |
| >21 days after first dose | 0.53 (0.41, 0.70) | 46.5% (30.4, 58.9) |
| **Fully adjusted^** |  |  |
| Unvaccinated | Reference | Reference |
| 0-13 days after first dose | 0.88 (0.56, 1.36) | 12.2% (-36.4, 43.5) |
| 14-21 days after first dose | 1.00 (0.63, 1.60) | -0.3% (-59.7, 37.0) |
| >21 days after first dose | 0.54 (0.41, 0.71) | 46.5% (29.4, 59.5) |

\* Adjusted by day of the year of RT-PCR testing (restricted cubic spline); ^ Adjusted by age (restricted cubic spline), sex, cardiovascular disease, respiratory disease, obesity, diabetes mellitus, immunosuppressed status (includes cancer), liver disease, occupation, region of residence, self-reported race, reason of testing, and day of the year of RT-PCR testing using a restricted cubic spline.

**eTable 5.** Vaccine effectiveness against asymptomatic COVID-19. (sensitivity analysis)

|  | **Symptomatic**<br>**(n = 4,951 tests)** |  |
| --- | --- | --- |
|  | **OR (95% CI)** | **VE (95% CI)** |
| **Adjusted by time of pandemic\*** |  |  |
| Unvaccinated | Reference | Reference |
| 0-13 days after first dose | 0.97 (0.41, 2.28) | 3.3% (-127.8, 59.0) |
| 14-21 days after first dose | 1.30 (0.50, 3.37) | -30.1% (-237.5, 49.8) |
| >21 days after first dose | 0.62 (0.31, 1.23) | 38.0% (-22.6, 68.7) |
| **Fully adjusted^** |  |  |
| Unvaccinated | Reference | Reference |
| 0-13 days after first dose | 1.01 (0.42, 2.42) | -1.1% (-142.3, 57.8) |
| 14-21 days after first dose | 1.35 (0.51, 3.55) | -35.0% (-255.5, 48.7) |
| >21 days after first dose | 0.70 (0.34, 1.44) | 29.8% (-44.2, 65.8) |

\* Adjusted by day of the year of RT-PCR testing (restricted cubic spline); ^ Adjusted by age (restricted cubic spline), sex, cardiovascular disease, respiratory disease, obesity, diabetes mellitus, immunosuppressed status (includes cancer), liver disease, occupation, region of residence, self-reported race, reason of testing, and day of the year of RT-PCR testing using a restricted cubic spline.

